# Vaccine effectiveness of primary and booster COVID-19 vaccinations against SARS-CoV-2 infection in the Netherlands from 12 July 2021 to 6 June 2022: a prospective cohort study

**DOI:** 10.1101/2023.01.09.23284335

**Authors:** Anne J. Huiberts, Brechje de Gier, Christina E. Hoeve, Hester E. de Melker, Susan J.M. Hahné, Gerco den Hartog, Diederick E. Grobbee, Janneke H.H.M. van de Wijgert, Susan van den Hof, Mirjam J. Knol

## Abstract

**Introduction:** Monitoring of COVID-19 vaccine effectiveness (VE) is needed to inform vaccine policy. We estimated VE of primary vaccination, and first and second booster vaccination, against SARS-CoV-2 infection overall, and in four risk groups defined by age and medical risk condition, in the Delta and Omicron BA.1/BA.2 periods.

**Methods:** VASCO is an ongoing prospective cohort study among vaccinated and unvaccinated Dutch adults. The primary endpoint was a self-reported positive SARS-CoV-2 test during 12 July 2021-6 June 2022. Participants with a prior SARS-CoV-2 infection, based on a positive test or serology, were excluded. We used Cox proportional hazard models with vaccination status as time-varying exposure and adjustment for age, sex, educational level, and medical risk condition. We stratified by Delta and Omicron BA.1/BA.2 periods, risk group, and time since vaccination.

**Results:** 37,170 participants (mean age 57 years) were included. In the Delta period, VE <6 weeks after primary vaccination was 80% (95%CI 69-87) and decreased to 71% (65-77) after 6 months. VE increased to 96% (86-99) shortly after the first booster vaccination. In the Omicron period these estimates were 46% (22-63), 25% (8-39) and 57% (52-62), respectively. VE was 50% (34-62) <6 weeks after a second booster vaccination in participants aged ≥60 years. For the Omicron period, an interaction term between vaccination status and risk group significantly improved the model (p<0.001), with generally lower VEs for those with a medical risk condition.

**Conclusions:** Our results show the benefit of booster vaccinations against infection, also in risk groups, although the additional protection wanes quite rapidly.

**Summary:** This prospective cohort study contributes to vaccine policy of COVID-19 by showing the benefit of booster vaccination in preventing SARS-CoV-2 infections, also in risk groups in which protection was generally lower, although the additional protection was rather short-lived.

## Introduction

After implementation of a vaccination program, real world vaccine effectiveness should be monitored to inform further vaccination policy [1]. The COVID-19 vaccination program in the Netherlands started on 6 January 2021. By 18 April 2021, four different COVID-19 vaccines had been approved and were used in the initial vaccination programme: Comirnaty (BNT162b2; BioNTech/Pfizer, Mainz, Germany/New York, United States (US)), Spikevax (mRNA-1273, Moderna, Cambridge, US), Vaxzevria (ChAdOx1-S; AstraZeneca, Cambridge, United Kingdom), and Jcovden (Ad26.COV2-S (recombinant), Janssen-Cilag International NV, Beerse, Belgium). Different vaccines were recommended and administered in varying age groups [2, 3]. The first booster campaign for adults was initiated on 18 November 2021, prioritizing health care workers and those ≥60 years. By June 2022, a primary series coverage of 83% and a booster vaccination coverage of 64% of the Dutch population ≥18 years had been reached [4]. From 4 March 2022, a second booster vaccination was offered to adults ≥60 years, and uptake was relatively low (44% by June 2022) [5].

Since the start of the vaccination programme, various new SARS-CoV-2 variants of concern emerged, including the Delta (B.1.617.2) and Omicron (B.1.1.529) variants. The Delta variant was first detected in the Netherlands in April 2021 and replaced the Alpha variant as the dominant strain in July 2021 [6]. The Omicron variant was first detected in late November 2021 and caused 90% of the infections six weeks later.

As in other countries, nationwide COVID-19 surveillance data in the Netherlands including testing and contact tracing data have been used to monitor and evaluate VE against SARS-CoV-2 infections [7-9]. The advantages of using national surveillance data are the large sample size and data availability in real-time. The disadvantages are dependence on testing infrastructure and testing behaviour. For example, the Dutch government scaled down free-of-charge testing at community test centers from 11 April 2022 onwards; the general public was encouraged to self-test when having symptoms from that date onwards.

The VAccine Study COvid-19 (VASCO) is a large population-based prospective cohort study that was initiated during the COVID-19 vaccination roll-out in the Netherlands enabled us to study vaccine-effectiveness irrespective of available registration data [10]. Both vaccinated and unvaccinated Dutch adults are followed for a five-year period during which extensive data, including demographics, vaccination data, and positive (self-)tests, are being collected from participants using regular online questionnaires.

Here we report on the VE of primary vaccination by any of the four available COVID-19 vaccines, as well as first and second booster vaccination, against self-reported SARS-CoV-2 infection by time since vaccination and in four subpopulations defined by age and medical risk condition, during 12 July 2021 to 6 June 2022, the period in which the Delta and Omicron BA.1 and BA.2 variants were sequentially dominant.

## Methods

### Study design and study population

VASCO is an ongoing population-based prospective cohort study with five-year follow-up [10]. The study was initiated during the roll-out of the COVID-19 vaccination program in the Netherlands. Between 3 May 2021 and 15 December 2021, 45,552 community-dwelling adults aged 18 to 85 years were included. Participants had to be able to understand Dutch, as all study materials were written in Dutch, and were included irrespective of their COVID-19 vaccination status or intention to get vaccinated. Participants were asked to complete monthly online questionnaires in the first year, and three-monthly online questionnaires in years 2-5, including questions on sociodemographic factors, health status, COVID-19 vaccination, SARS-CoV-2-related symptoms, testing results, and test intention. At inclusion, 6 and 12 months after inclusion, and one month after primary vaccination, participants were asked to take a self-collected fingerprick blood sample at home. Samples were tested for SARS-CoV-2 antibodies by Elecsys Anti-SARS-CoV-2 immunoassay (Roche Diagnostics, Vienna, Austria) using the NIBSC 20/136 WHO standard for quantification. In the current analysis, serology data were used to identify participants who had had a SARS-CoV-2 infection prior to the study period by determining the presence of immunoglobulin (Ig) antibodies against the SARS-CoV-2 nucleocapsid protein (anti-N). Written informed consent was obtained from all participants prior to enrollment into the study. The VASCO study is conducted in accordance with the principles of the Declaration of Helsinki and the study protocol was approved by the not-for-profit independent Medical Ethics Committee of the *Stichting Beoordeling Ethiek Biomedisch Onderzoek* (BEBO), Assen, the Netherlands).

### Vaccination status

Self-reported vaccination data were linked to vaccination data registered in the Dutch national COVID-19 vaccination Information and Monitoring System (CIMS) [3]. Vaccination data from the CIMS registry were considered the primary source, except when the participant did not provide informed consent for vaccination registration in CIMS or for linking study and CIMS data. If CIMS and/or self-reported data were incomplete, data from both sources were combined (see **Additional file 1 and Table S1** for a detailed description). Vaccination status was categorized as unvaccinated (no vaccination received), primary vaccination series received (one dose of Jcovden 28+ days ago, or two doses of Vaxzevria, Comirnaty or Spikevax 14+ days ago), primary vaccination series and one booster received (primary vaccination series + one additional dose 7+ days ago), or primary vaccination series and two boosters received (primary vaccination series + two additional doses 7+ days ago) [2, 3]. For individuals with a severe immune deficiency primary vaccination consisted of three doses. Therefore, a third dose administered before the start of the general public booster campaign (18 November 2021) was considered an additional primary series vaccination and not a booster vaccination. A second booster vaccination in the spring of 2022 was only available for individuals aged 60 years and above and some highly vulnerable groups. The 7, 14 or 28 person-days between vaccine administration and obtained vaccination status were excluded because we assumed that immunity was not yet fully established. Participants were excluded if they reported to have received more doses than possible according to the Dutch vaccination strategy [2].

### SARS-CoV-2 infections

The primary endpoint was a self-reported positive SARS-CoV-2 test. Participants were asked to notify all positive SARS-CoV-2 tests via the study website or app (either a test by a community testing center free-of-charge, a test at a commercial test center, or a self-administered antigen-test). Community testing was scaled down from 11 April 2022 onwards. To facilitate testing after that date in case of symptoms associated with COVID-19 and/or contact with a person infected with SARS-CoV-2, the study team provided self-tests to participants from May 2022 onwards. Participants could report positive tests in real time, and in addition, each scheduled follow-up questionnaire contained questions regarding recent positive SARS-CoV-2 tests. Reported infections were considered Delta infections if the positive test date was between 12 July 2021 and 19 December 2021, the period in which >90% of the cases was caused by the Delta variant [11]. Reported positive tests from 10 January 2022 until 6 June 2022 were attributed to the Omicron BA.1 or BA.2 variant. Participants who had reported a positive test or tested positive for N-antibodies prior to start of follow-up in the current analysis were excluded from the analysis in order to estimate effects of vaccination only.

### Covariates

Sociodemographic data such as age and sex were collected at baseline and during follow-up. Educational level was classified as low (no education or primary education), intermediate (secondary school or vocational training), or high (bachelor’s degree, university). A medical risk condition was present when a participant reported to have one or more of the following conditions: diabetes mellitus, lung disease or asthma, asplenia, cardiovascular disease, immune deficiency, cancer (currently untreated but treated in the past, currently treated, untreated), liver disease, neurological disease, renal disease, organ or bone marrow transplantation. Four risk groups were defined by age (18-59 and 60-85 years) and presence of a medical risk condition (present or absent).

### Statistical analyses

Data were inspected using descriptive statistics and graphical displays. Cox proportional hazard models were used to estimate VE of primary series, and first and second booster vaccination against SARS-CoV-2 infection in the Delta and Omicron BA.1/BA.2 period. Vaccination status was included as a time-varying exposure. Participants entered the study at the start of the study period (12 July 2021) or the date of completion of the baseline questionnaire, if they became a participant later than 12 July 2021. Participants were followed until the date of the first reported positive test. If no positive test was reported, participants were followed until the most recent questionnaire completion date plus the median time between follow-up questionnaires for that participant (to include only person-time in which participants were assumed to be active), or the end date of the study period (6 June 2022), whichever came first. Median time between follow-up questionnaires was determined per person and separately for the first year in which participants received monthly questionnaires, and after year 1 when they received questionnaires every three months. Calendar time was used as the underlying timescale for the Cox regression. This effectively means that at each date participants with different vaccination statuses were compared, thereby adjusting for factors changing over time during the pandemic, i.e. infection pressure, and the number of vaccinated persons in the population. Potential violation of assumptions regarding proportional hazards was checked using graphical diagnostics based on the scaled Schoenfeld residuals.

Models were stratified by Delta and Omicron periods and by time since start of the vaccination status in 6-week intervals. Analyses were first adjusted for sex, educational level, and age group, and then additionally for the presence of a medical risk condition. Age group and the presence of a medical risk condition were included as time-varying confounders. Risk group membership based on age (18-59 and 60-85 years) and medical risk condition (present or absent) was examined as a potential effect modifier by extending the model with an interaction term and by stratified analysis. As sensitivity analyses, analyses were repeated in two specific subpopulation. The first sensitivity analysis was done in participants who reported to (almost) always test for SARS-CoV-2 infection in case of SARS-CoV-2 related symptoms. Secondly, the analysis was repeated in participants who had received only Comirnaty vaccine doses versus unvaccinated participants. We also present VE estimates of the primary vaccination series stratified by vaccine product (Comirnaty, Spikevax, Vaxzevria and Jcovden) and VE estimates of first booster vaccination stratified by vaccine product of the booster (Comirnaty or Spikevax) and primary vaccination series (mRNA vaccine or Vaxzevria).

Vaccine effectiveness was calculated as 100% x (1 – hazard ratio). All statistical analyses were performed in statistical package R version 4.1.3, using packages Epi and survival.

## Results

### Study population

Of the 45,049 VASCO participants participating (partly) during the study period, 121 participants were excluded because of having had a first and/or second booster vaccination before the start of that particular booster campaign. Additionally, 40 participants were excluded because of missing covariates and 255 participants did not add person-time to one of the studied vaccination status strata, e.g. entered the study when already having received a first vaccination but never completed primary vaccination series (not applicable to Jcovden). Of the 44,633 remaining participants, 6,826 participants (15.3%) reported to have had a positive SARS-CoV-2 test prior to start of follow-up in the current analysis. Additionally, 991 (2.2%) tested positive for SARS-CoV-2 anti-N antibodies prior to start of follow-up. Consequently, 36,816 participants were included in the analyses (**Table 1**). The age of the participants at inclusion ranged between 18 and 85 years, with a median age of 61 years. More women (62%) than men were included, and 57% of the participants was highly educated. At the start of the study period, 12,152 participants were included in the study, of which 11,908 (98.0%) had completed their primary vaccination series, and 244 (2.0%) were unvaccinated (**Additional file 1, Figure S1**). At the start of the Omicron period, the cohort consisted of 27,646 active participants. Of those, 3,802 (13.8%) participants had only completed their primary vaccination series, 23,352 (84.5%) participants had additionally received a first booster vaccination, and 492 (1.8%) participants were unvaccinated. Of all participants that contributed vaccinated person-weeks during the study period (n=36,109), the first vaccine dose was most often Comirnaty (41.5%). Other first vaccination products were Vaxzevria (33.7%), Spikevax (13.0%), Jcovden (9.9%), other (0.01%) or unknown (0.1%).

**Table 1.** Baseline characteristics of participants included in analysis

|  | **Total**<br>**(n=36,816)** | **18-59 years**<br>**(n=16,575)** | **60-85 years**<br>**(n=20,241)** |
| --- | --- | --- | --- |
| Sex (%) |  |  |  |
| Male | 13,874 (37.7) | 4,633 (28.0) | 9,241 (45.7) |
| Female | 22,922 (62.3) | 11,923 (71.9) | 10,999 (54.3) |
| Other | 20 (0.1) | 19 (0.1) | 1 (0.0) |
| Median age (years; IQR) | 61 (15) | 48 (17) | 65 (7) |
| Medical risk condition<sup>a</sup> at inclusion, yes (%) | 11,078 (30.1) | 3,307 (20.0) | 7,771 (38.4) |
| Cardiovascular disease | 6,612 (18.0) | 1,322 (8.0) | 5,290 (26.1) |
| Lung disease or asthma | 2,852 (7.7) | 1,289 (7.8) | 1,563 (7.7) |
| Diabetes mellitus | 1,820 (4.9) | 393 (2.4) | 1,427 (7.1) |
| Immune deficiency | 697 (1.9) | 336 (2.0) | 361 (1.8) |
| Educational level<sup>b</sup> (%) |  |  |  |
| Low | 5,151 (14.0) | 1,105 (6.7) | 4,046 (20.0) |
| Intermediate | 10,328 (28.1) | 4,976 (30.0) | 5,352 (26.4) |
| High | 21,119 (57.4) | 10,441 (63.0) | 10,678 (52.8) |
| Other | 218 (0.6) | 53 (0.3) | 165 (0.8) |

<sup>a</sup> Medical risk condition: one or more of following conditions: diabetes mellitus, lung disease or asthma, asplenia, cardiovascular disease, immune deficiency, cancer (currently untreated, currently treated, untreated), liver disease, neurological disease, renal disease, organ or bone marrow transplantation. Four most frequent conditions are presented here.
<sup>b</sup> Educational level was classified as low (no education or primary education), intermediate (secondary school or vocational training), or high (bachelor's degree, university).

Participants had a median follow-up time of 27.7 person weeks. This was relatively short as participants were included over a period of 7 months and were censored after a reported SARS-CoV-2 test. During a total of 1,032,976 person-weeks of follow-up, 13,756 first SARS-CoV-2 infections were reported corresponding with an infection rate of 13.3 infections per 1,000 person-weeks. Reported positive tests were often a PCR-test (72.7%) or antigen-test (can be self-administered) (26.1%), with the share of antigen-tests increasing sharply during the Omicron period (**Additional file 1, Figure S2**). The largest proportion of reported infections (12,129, 88.2%) occurred during the Omicron BA.1/BA.2 period (**Figure 1**). Infection rates were higher during person-weeks for unvaccinated compared to person-weeks for vaccinated (**Figure 1, Table 2**).

**Figure 1.**
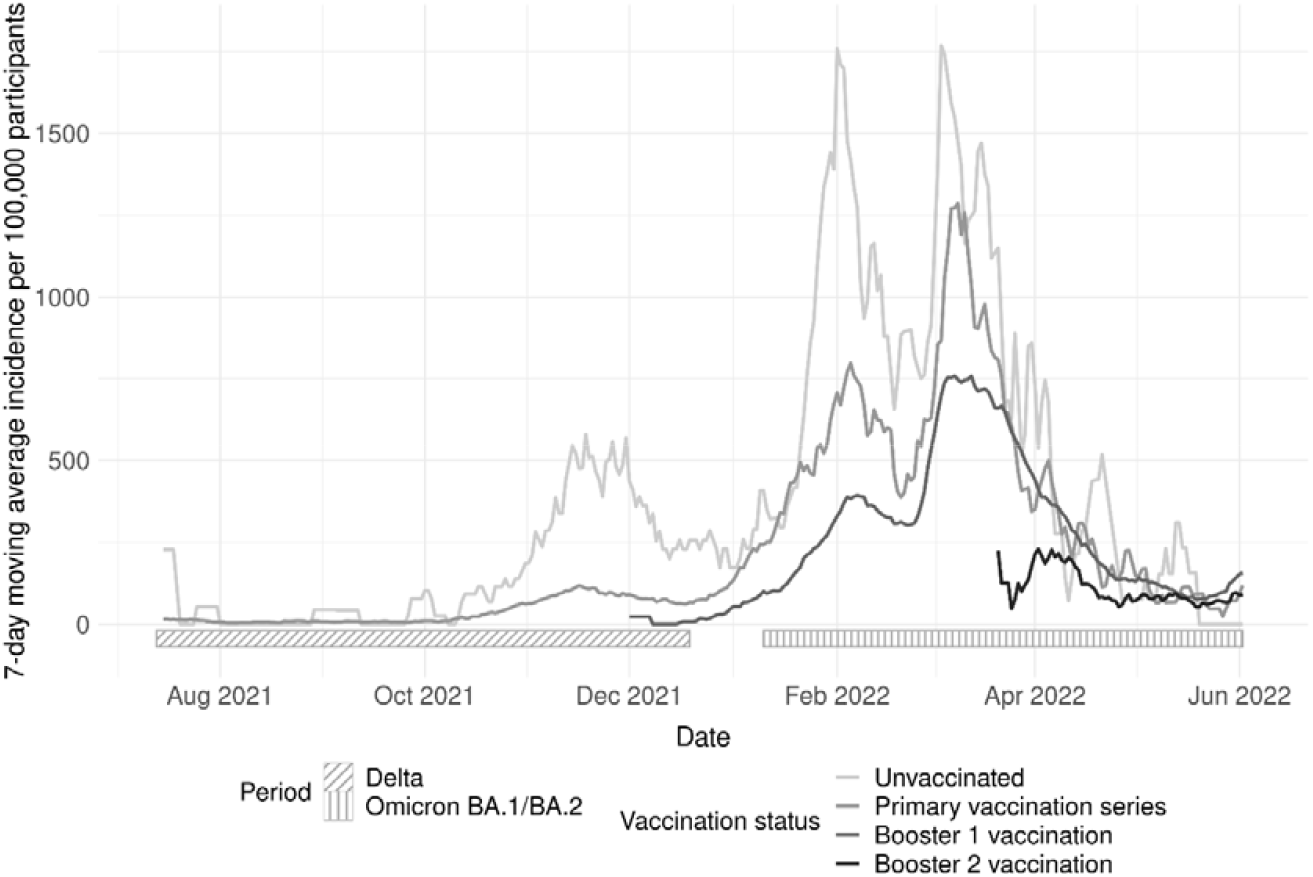
7-days moving average of number of infections reported per 100,000 VASCO participants by vaccination status from 12 July 2021 to 6 June 2022

**Table 2.**
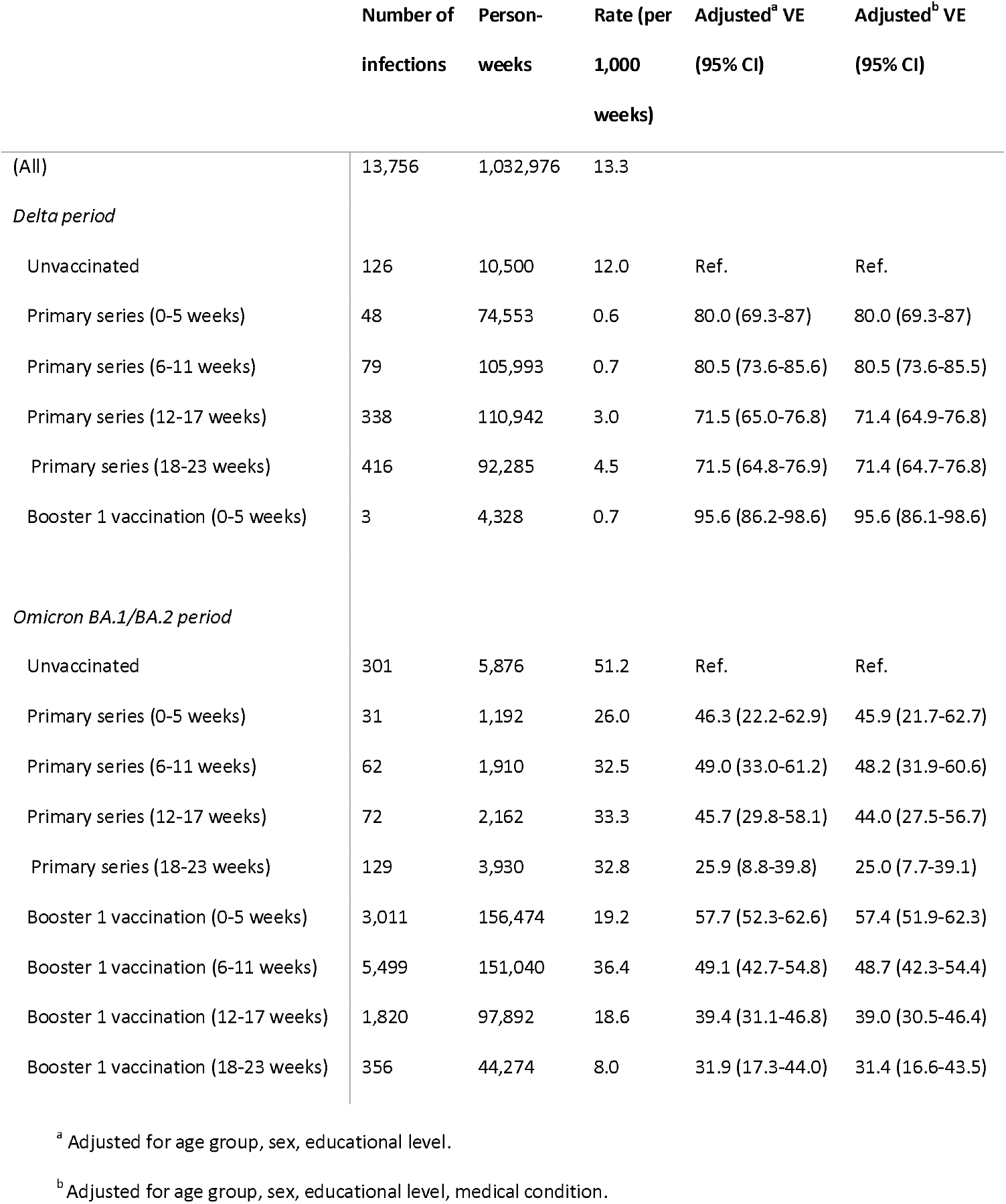
Vaccine effectiveness per vaccination status stratified by Delta and Omicron BA.1/BA.2 period from 12 July 2021 to 6 June 2022

|  | Number of<br>infections | Person-weeks | Rate (per<br>1,000<br>weeks) | Adjusted<sup>a</sup> VE<br>(95% CI) | Adjusted<sup>b</sup> VE<br>(95% CI) |
| --- | --- | --- | --- | --- | --- |
| (All) | 13,756 | 1,032,976 | 13.3 |  |  |
| *Delta period* |  |  |  |  |  |
| Unvaccinated | 126 | 10,500 | 12.0 | Ref. | Ref. |
| Primary series (0-5 weeks) | 48 | 74,553 | 0.6 | 80.0 (69.3-87) | 80.0 (69.3-87) |
| Primary series (6-11 weeks) | 79 | 105,993 | 0.7 | 80.5 (73.6-85.6) | 80.5 (73.6-85.5) |
| Primary series (12-17 weeks) | 338 | 110,942 | 3.0 | 71.5 (65.0-76.8) | 71.4 (64.9-76.8) |
| Primary series (18-23 weeks) | 416 | 92,285 | 4.5 | 71.5 (64.8-76.9) | 71.4 (64.7-76.8) |
| Booster 1 vaccination (0-5 weeks) | 3 | 4,328 | 0.7 | 95.6 (86.2-98.6) | 95.6 (86.1-98.6) |
| *Omicron BA.1/BA.2 period* |  |  |  |  |  |
| Unvaccinated | 301 | 5,876 | 51.2 | Ref. | Ref. |
| Primary series (0-5 weeks) | 31 | 1,192 | 26.0 | 46.3 (22.2-62.9) | 45.9 (21.7-62.7) |
| Primary series (6-11 weeks) | 62 | 1,910 | 32.5 | 49.0 (33.0-61.2) | 48.2 (31.9-60.6) |
| Primary series (12-17 weeks) | 72 | 2,162 | 33.3 | 45.7 (29.8-58.1) | 44.0 (27.5-56.7) |
| Primary series (18-23 weeks) | 129 | 3,930 | 32.8 | 25.9 (8.8-39.8) | 25.0 (7.7-39.1) |
| Booster 1 vaccination (0-5 weeks) | 3,011 | 156,474 | 19.2 | 57.7 (52.3-62.6) | 57.4 (51.9-62.3) |
| Booster 1 vaccination (6-11 weeks) | 5,499 | 151,040 | 36.4 | 49.1 (42.7-54.8) | 48.7 (42.3-54.4) |
| Booster 1 vaccination (12-17 weeks) | 1,820 | 97,892 | 18.6 | 39.4 (31.1-46.8) | 39.0 (30.5-46.4) |
| Booster 1 vaccination (18-23 weeks) | 356 | 44,274 | 8.0 | 31.9 (17.3-44.0) | 31.4 (16.6-43.5) |

<sup>a</sup> Adjusted for age group, sex, educational level.
<sup>b</sup> Adjusted for age group, sex, educational level, medical condition.

### Vaccine effectiveness

Fully adjusted VE in the Delta period was estimated to be 80% (95%CI 69.3 – 87.0) <6 weeks after completing the primary series, counting from the start of the vaccination status not vaccine administration (**Figure 2**). This decreased to 71% (95%CI 64.7 – 76.8) 19 – 24 weeks after completion of the primary vaccination series. VE increased again to 96% (95%CI 86.1 – 98.6) <6 weeks after the booster vaccination. VE estimates for the Omicron period were substantially lower compared to those in the Delta period. VE in the first 6 weeks after completing the primary vaccination series was estimated to be 46% (95%CI 21.7 – 62.7) and decreased to 25% (95%CI 7.7 – 39.1) 18-23 weeks after completion of the primary vaccination series. VE increased to 57% (95%CI 51.9 – 62.3) <6 weeks after booster vaccination and decreased to 31% (95%CI 16.6 – 43.5) at 18-23 weeks. For participants of 60 years and older, the VE against Omicron infection within 6 weeks after the second booster vaccination was 50% (95% CI 34.0 – 62.1) (**Figure 3; Additional file 1, Table S2**). Delta VE estimates of the models with and without having a medical risk condition (yes/no) as confounder were comparable (**Table 2**). VE estimates for Omicron were slightly lower when additionally adjusting for the presence of a medical risk condition.

**Figure 2.**
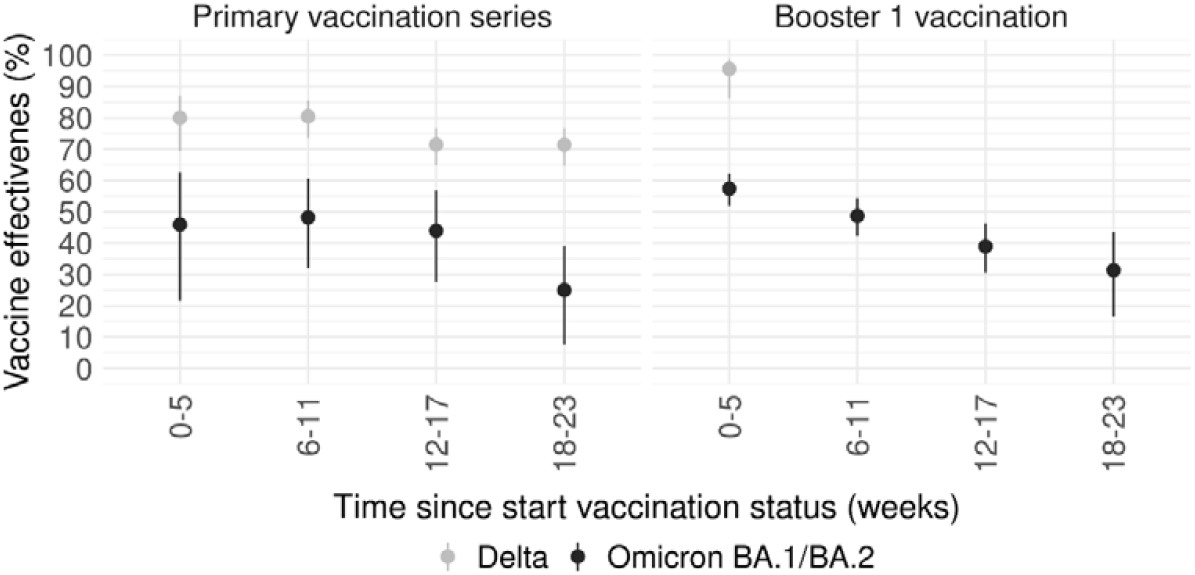
Vaccine effectiveness^a^ for primary vaccination series and first booster vaccination in Delta and Omicron BA.1/BA.2 period from 12 July 2021 to 6 June 2022 ^a^ Adjusted for age group, sex, educational level, medical condition.

**Figure 3.**
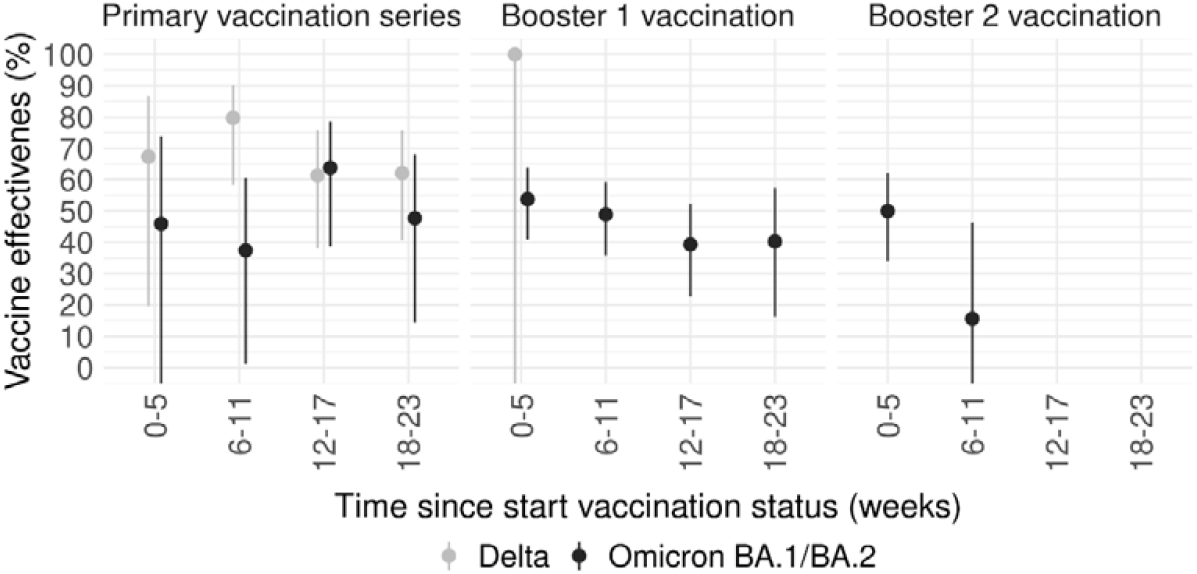
Vaccine effectiveness^a^ for primary vaccination series, first booster and second booster vaccination in Delta and Omicron BA.1/BA.2 period in participants aged ≥60 years from 12 July 2021 to 6 June 2022 ^a^ VE was not reported when number of person-weeks <500; Adjusted for age group, sex, educational level, medical condition.

VE estimates showed a similar pattern in the sensitivity analysis restricted to participants with a high intention to test in case of symptoms (n=26,520, median age = 61) except that VE estimates for Omicron infection were higher in this specific population (**Additional file 1, Figure S3**). The higher estimates in this sensitivity analysis were in line with a higher intention to test in vaccinated participants (**Additional file 1, Figure S4**).

In the sensitivity analysis restricted to participants who only received Comirnaty vaccine doses (as primary series and as booster(s) if booster(s) were received) (n=14,652 (39.8% of full analysis population), median age = 60), VE estimates for the Delta period were comparable to the VE estimates of the complete study population (**Additional file 1, Table S3**). VE against Delta infection decreased from 81% (95%CI 69.3 – 88.6) within 6 weeks after completion of primary series to 72% (95%CI 64.9 – 78.3) 18-23 weeks after completion and increased to 96% (95%CI 67.7 – 99.4) within 6 weeks after booster vaccination. For the Omicron period, VE estimates for the booster vaccination were slightly but consistently lower in the Comirnaty subpopulation compared to the total study population. VE decreased from 51% (95%CI 43.6 – 57.8) within 6 weeks after booster vaccination to 11% (95%CI -27.0 – 36.9) 18-23 weeks after the booster. VE estimates stratified by vaccine product of the primary series and first booster vaccination are given in **Additional file 1, Table S5 and S6**. Generally, estimates were higher for Spikevax as primary series and lower for Vaxzevria and Jcovden compared with Comirnaty. For booster vaccination, estimates for Spikevax as booster were generally higher compared with Comirnaty as booster, irrespective of the vaccine product of the primary series.

For the Delta period, models with and without an interaction term between vaccination status and risk group did not differ significantly. For the Omicron period, the interaction term did significantly improve the model (p<0.001). The interaction term was significant between at least two risk groups for all periods after booster vaccination (**Additional file 1, Table S4**). When stratifying the model according to risk group, VE of booster vaccination in the Omicron period was lower among participants with a medical condition as compared to those without (**Figure 4**). Number of infections and person-weeks in unvaccinated persons with medical risk condition were relatively small, resulting in large confidence intervals around the VE.

**Figure 4.**
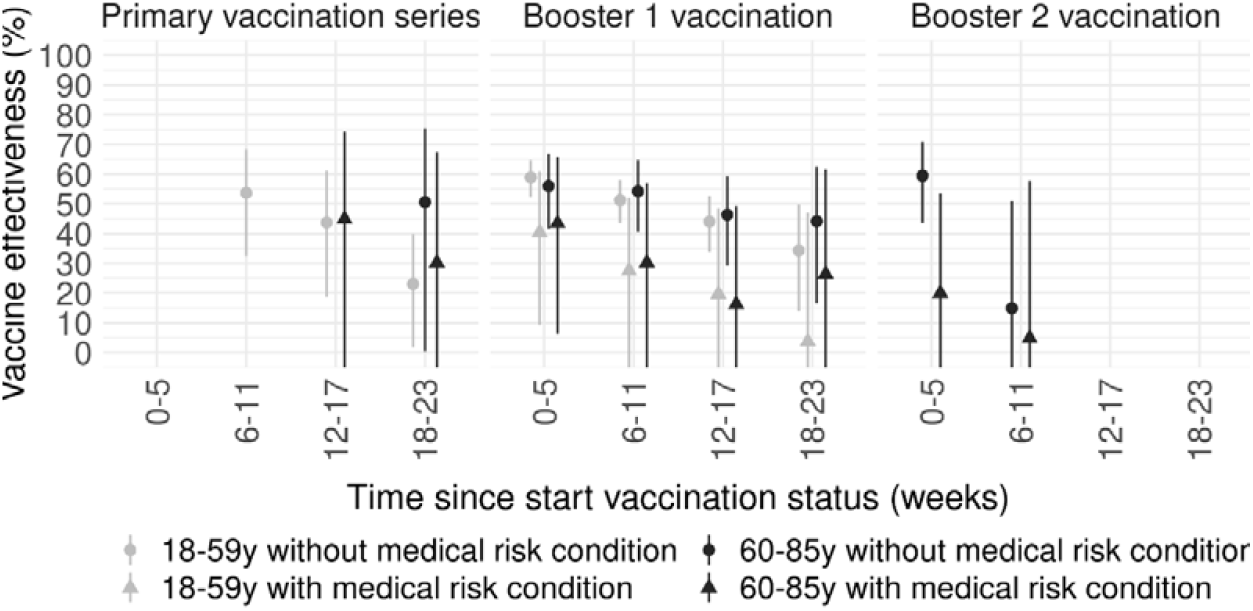
Vaccine effectiveness^a^ for primary vaccination series, booster and second booster vaccination per risk group in the Omicron BA.1/BA.2 period from 10 January 2022 to 6 June 2022 ^a^ VE was not reported when number of person-weeks <500; Adjusted for age group, sex, educational level.

## Discussion

We evaluated the effectiveness of COVID-19 vaccines against Delta and Omicron BA.1/BA.2 SARS-CoV-2 infection in a real-world setting, overall and in four risk groups based on age and presence of medical risk condition. Compared to unvaccinated individuals, having completed the primary vaccination series was associated with protection against Delta and Omicron BA.1/BA.2 SARS-CoV-2 infection. However, the protection against infection with the Omicron BA.1/BA.2 variants was markedly lower compared to protection against infection with the Delta variant. VE decreased over time after completing the primary vaccination series, but increased again after receiving a first booster vaccination, also in risk groups. In those aged 60 years and older, VE increased again after receiving a second booster. VE of booster vaccinations also decreased over time since vaccination. Our data showed that unvaccinated participants had a lower intention to test if having symptoms compared to vaccinated participants. Indeed, when restricting our analysis to participants with high intention to test, VE against Omicron infection was higher. Despite large confidence intervals, VE against Omicron BA.1/BA.2 SARS-CoV-2 infection appeared lower among participants with a medical risk condition compared to participants without a medical risk condition, visible both in younger and older individuals. Our estimates concern effects of vaccination only, as prior infections were excluded.

Our study results are in line with national [7] and international surveillance data [12-15], showing a higher VE against Delta infection as compared to Omicron infection, resulting from considerable immune escape by the Omicron variant [16, 17]. Reported estimates for VE shortly after completion of a primary series range between 78% and 91% against Delta infection and 40% and 66% against Omicron infection are consistent with our findings of 80% and 46%, respectively. VE estimates of booster vaccination against Delta (96%) and Omicron infection (57%) were consistent with those found using surveillance data (86%-99% and 56%-72%, respectively) [7, 12-15]. Similar to our findings, other studies have shown waning of the effectiveness of both primary and booster vaccination [12-14]. Data on VE of second booster vaccination with unvaccinated as reference group is scarce. One preprint reported a VE against BA.2 infection in adults of 64% (95%CI 50.7 – 74.2) 14-30 days after fourth dose, which decreased to 51% (95%CI 35.5 – 63.0) 31-90 days after the fourth dose [18]. Our estimates were slightly lower (50% after 0-5 weeks and 16% after 6-11 weeks) but were based on data of adults aged 60 years and older only.

Only two other prospective cohort studies have reported VEs against Delta infection [19, 20]. In both studies, nose and/or throat swabs for PCR testing were regularly collected irrespective of having symptoms, allowing detection of symptomatic as well as asymptomatic infections. VE of Comirnaty (BioNTech/Pfizer) primary vaccination series in the Delta period reported in the ONS CIS study decreased from 85% at 14 days after second dose to 75% at 90 days [20]. Results of our sensitivity analysis in participants who had only received Comirnaty vaccine doses were consistent with the ONS CIS Comirnaty estimates (81% at 0-5 weeks and 79% at 6-11 weeks after primary series). The VE estimate in the HEROES-RECOVER study was lower (66%, 95%CI 26-84), but time since vaccination was not taken into account and the study population consisted of health care workers only with likely high exposure [19]. The ONS CIS study further showed that VE of Vaxzevria (AstraZeneca) primary vaccination series was considerably lower than for Comirnaty (68% at 14 days and 61% at 90 days), which was consistent with our results. Our results showed that VE of Comirnaty booster vaccination was lower as compared to VE of Spikevax booster vaccination. This is consistent with literature showing higher antibody levels after a Spikevax booster [21].

There is limited data on VE against infection in medical risk populations. One test-negative case-control study evaluated three-dose VE against infection in immunocompromised and immunocompetent individuals [13]. They found a significant interaction between immunocompromised status and vaccination status in both Delta and Omicron periods. In both periods, stratified analysis showed a lower VE in immunocompromised individuals (Delta: 70.6%, 95%CI 31.0 – 87.5 ; Omicron: 29.4%, 95%CI 0.3 – 50.0) as compared to immunocompetent individuals (Delta: 93.7%, 95%CI 92.2 – 94.9; Omicron: 70.5%, 95% CI 68.6 – 72.4). Differences between the groups were larger than the differences we observed, yet our definition of medical risk was broader than immunocompromised individuals only. An Israelian historic cohort study showed lower VE of two doses against infection in both individuals with diabetes and cardiovascular disease (82%, 95% CI 62 – 92) and immunocompromised individuals (71%, 95% CI 37 – 87) as compared to overall (92%, 95%CI 83 – 96) [22]. Taking into account increased risk for severe COVID-19 outcomes [23], our results support the Dutch vaccination strategy to recommend booster vaccination for high risk groups.

This study has several strengths. In this cohort study we were able to adjust for (time-varying) confounders using extensive data from monthly questionnaires. Also, serological data enabled us to exclude participants with prior unreported SARS-CoV-2 infections. A recent study by Kahn et al emphasized the added value of serological testing to exclude participants with prior infection [24]. Also we were able to include self-administered antigen tests, freely available to participants, as outcome so we were not dependent on the testing infrastructure and we facilitated the use of self-tests by providing those to the participants. Further, the questionnaire on test behaviour allowed for an analysis restricted to participants with a consistently high intention to test in case of symptoms.

Some limitations need to be discussed. Although the Cox proportional hazards models were adjusted for potential confounders, differences in (time-varying) factors between vaccinated and unvaccinated participants which may impact infection exposure can still confound the results. These include differences in test frequency and differences in exposure through behaviour or adherence to COVID-19 guidelines. Vaccinated individuals in our cohort had higher intention to test when symptoms occurred than unvaccinated individuals, possibly they are more health-conscious. Still, vaccination may have reduced testing if breakthrough infections are more often mild or asymptomatic. In other contexts, vaccinated individuals may test less frequently, if vaccination induces a sense of security and people are less worried, or if public health authorities request more frequent testing of unvaccinated individuals. From the start of the study period until the end of March 2022, when most COVID-19 interventions were lifted, use of the corona check app was in place, which for unvaccinated individuals required a negative PCR test, for example to enter restaurants and clubs. These behavioural factors might have resulted in either an underestimation or overestimation of the VE. Furthermore, vaccinated individuals may become more heavily exposed to the virus, if they feel more safe to attend (high-risk) exposure activities [25]. Even though it is suggested that there is little change in behaviour early after vaccination [26] and a recent study showed that differences in chance of SARS-CoV-2 exposure due to behaviour did not relevantly confound VE estimates in a test-negative setting [27], this phenomenon might decrease the benefit of vaccination [28].

Overall, our results show that VE was lower against Omicron infection than Delta infection, and both first and second booster vaccination increased waned effectiveness again, although the additional protection was rather short-lived. Importantly, this booster effect was also seen among risk groups but protection of vaccination against Omicron infection was consistently lower among risk groups. Thus, our data shows the benefit of booster vaccination in preventing SARS-CoV-2 infections, also in risk groups.

## Data Availability

All data produced in the present study are available in aggregated and anonymized form upon reasonable request to the authors.

## List of abbreviations

BEBO: Beoordeling Ethiek Biomedisch Onderzoek
CIMS: COVID-19 vaccination Information and Monitoring System
Ig: immunoglobulin
N: nucleocapsid protein
RIVM: National Institute of Public Health and the Environment
VASCO: VAccine Study COvid-19
VE: vaccine effectiveness
WHO: World Health Organization

## References

1. Hahné S, Bollaerts K, Farrington P. Vaccination Programmes: Epidemiology, Monitoring, Evaluation. 1st ed: Routledge, 2021.

2. Pluijmaekers A, de Melker H. The National Immunisation Programme in the Netherlands. Surveillance and developments in 2021-2022. Het Rijksvaccinatieprogramma in Nederland Surveillance en ontwikkelingen in 2021-2022: Rijksinstituut voor Volksgezondheid en Milieu RIVM, 2022.

3. Valk A, van Meijeren D, Smorenburg N, et al. Vaccinatiegraad COVID-19 vaccinatie Nederland, 2021. COVID-19 vaccination coverage in the Netherlands in 2021: Rijksinstituut voor Volksgezondheid en Milieu RIVM, 2022.

4. Rijksoverheid. Coronadashboard: De actuele situatie in Nederland. Available at: https://coronadashboard.rijksoverheid.nl/. Accessed 22 March 2022.

5. Rijksinstituut voor Volksgezondheid en Milieu. Archief wekelijkse update vaccinatiecijfers 2022. Available at: https://www.rivm.nl/covid-19-vaccinatie/archief-wekelijkse-update-vaccinatiecijfers-2022. Accessed 24 June 2022.

6. RIVM COVID-19 epidemiologie en surveillance team. Effectiviteit van COVID-19-vaccinatie tegen SARS-CoV-2 infectie in de Delta periode, 2021 16 December 2021.

7. Andeweg SP, de Gier B, Eggink D, et al. Protection of COVID-19 vaccination and previous infection against Omicron BA.1, BA.2 and Delta SARS-CoV-2 infections. Nat Commun 2022; 13(1): 4738.

8. de Gier B, Andeweg S, Joosten R, et al. Vaccine effectiveness against SARS-CoV-2 transmission and infections among household and other close contacts of confirmed cases, the Netherlands, February to May 2021. Euro Surveill 2021; 26(31).

9. de Gier B, Andeweg S, Backer JA, et al. Vaccine effectiveness against SARS-CoV-2 transmission to household contacts during dominance of Delta variant (B.1.617.2), the Netherlands, August to September 2021. Euro Surveill 2021; 26(44).

10. Huiberts A, Kooijman M, Melker H, et al. Design and baseline description of an observational population-based cohort study on COVID-19 vaccine effectiveness in the Netherlands - The VAccine Study COvid-19 (VASCO). 2022.

11. Rijksinstituut voor Volksgezondheid en Milieu. Varianten van het coronavirus SARS-CoV-2. Available at: https://www.rivm.nl/coronavirus-covid-19/virus/varianten. Accessed 29 July 2022.

12. Suarez Castillo M, Khaoua H, Courtejoie N. Vaccine-induced and naturally-acquired protection against Omicron and Delta symptomatic infection and severe COVID-19 outcomes, France, December 2021 to January 2022. Eurosurveillance 2022; 27(16): 2200250.

13. Tseng HF, Ackerson BK, Luo Y, et al.Effectiveness of mRNA-1273 against SARS-CoV-2 Omicron and Delta variants. Nat Med 2022; 28(5): 1063–71.

14. Andrews N, Stowe J, Kirsebom F, et al. Covid-19 Vaccine Effectiveness against the Omicron (B.1.1.529) Variant. N Engl J Med 2022; 386(16): 1532–46.

15. Gram MA, Emborg H-D, Schelde AB, et al. Vaccine effectiveness against SARS-CoV-2 infection and COVID-19-related hospitalization with the Alpha, Delta and Omicron SARS-CoV-2 variants: a nationwide Danish cohort study. medRxiv 2022: 2022.04.20.22274061.

16. Yu J, Collier A-rY, Rowe M, et al. Neutralization of the SARS-CoV-2 Omicron BA.1 and BA.2 Variants. New England Journal of Medicine 2022; 386(16): 1579–80.

17. Netzl A, Tureli S, LeGresley E, Mühlemann B, Wilks SH, Smith DJ. Analysis of SARS-CoV-2 Omicron Neutralization Data up to 2021-12-22. bioRxiv 2022: 2021.12.31.474032.

18. Tseng HF, Ackerson BK, Bruxvoort KJ, et al. Effectiveness of mRNA-1273 against infection and COVID-19 hospitalization with SARS-CoV-2 Omicron subvariants: BA.1, BA.2, BA.2.12.1, BA.4, and BA.5. medRxiv 2022: 2022.09.30.22280573.

19. Fowlkes A, Gaglani M, Groover K, Thiese MS, Tyner H, Ellingson K. Effectiveness of COVID-19 Vaccines in Preventing SARS-CoV-2 Infection Among Frontline Workers Before and During B.1.617.2 (Delta) Variant Predominance — Eight U.S. Locations, December 2020–August 2021. MMWR Morb Mortal Wkly Rep 2021; 70: 1167–9.

20. Pouwels KB, Pritchard E, Matthews PC, et al. Effect of Delta variant on viral burden and vaccine effectiveness against new SARS-CoV-2 infections in the UK. Nat Med 2021; 27(12): 2127–35.

21. Wei J, Matthews PC, Stoesser N, et al. Correlates of protection against SARS-CoV-2 Omicron variant and anti-spike antibody responses after a third/booster vaccination or breakthrough infection in the UK general population. medRxiv 2022: 2022.11.29.22282916.

22. Chodick G, Tene L, Rotem RS, et al. The Effectiveness of the Two-Dose BNT162b2 Vaccine: Analysis of Real-World Data. Clin Infect Dis 2022; 74(3): 472–8.

23. Williamson EJ, Walker AJ, Bhaskaran K, et al. Factors associated with COVID-19-related death using OpenSAFELY. Nature 2020; 584(7821): 430–6.

24. Kahn R, Schrag SJ, Verani JR, Lipsitch M. Identifying and Alleviating Bias Due to Differential Depletion of Susceptible People in Postmarketing Evaluations of COVID-19 Vaccines. Am J Epidemiol 2022; 191(5): 800–11.

25. Ioannidis JPA. Factors influencing estimated effectiveness of COVID-19 vaccines in non-randomised studies. BMJ Evidence-Based Medicine 2022: bmjebm-2021-111901.

26. Goldszmidt R, Petherick A, Andrade EB, et al. Protective Behaviors Against COVID-19 by Individual Vaccination Status in 12 Countries During the Pandemic. JAMA Network Open 2021; 4(10): e2131137–e.

27. van Ewijk CE, Kooijman MN, Fanoy E, et al. COVID-19 vaccine effectiveness against SARS-CoV-2 infection during the Delta period, a nationwide study adjusting for chance of exposure, the Netherlands, July to December 2021. Euro Surveill 2022; 27(45).

28. Ioannidis JPA. Benefit of COVID-19 vaccination accounting for potential risk compensation. npj Vaccines 2021; 6(1): 99.

