## Additional file 1 for "Vaccine effectiveness of primary and booster COVID-19 vaccinations against SARS-CoV-2 infection in the Netherlands from 12 July 2021 to 6 June 2022: a prospective cohort study"

**Additional information combining CIMS and self-reported vaccination data**

*Cleaning* – Data was extracted from CIMS registry on 10 March 2022. Firstly, both self-reported (data of 4 July 2022) and registry vaccination data were cleaned. Vaccinations were removed if administered before the start of the vaccination roll out in the Netherlands (6 January 2021). Vaccinations reported in the beginning of 2022 with a vaccination date in the beginning of 2021 were assumed to be typing errors and were changed to the year 2022. In case of two vaccines administered within three weeks according to CIMS, the last vaccination was removed. If this was the case for self-reported vaccinations, both vaccinations were kept for comparison with CIMS but were numbered as the same dose.

*Comparing and combining* – The first five doses were compared between CIMS and self-reported data. In the beginning of the study, participants were asked to report dates of all received previous doses when reporting a new dose. Therefore, for self-reported data, there was often more than one date reported per dose. 2.4% of all participants did not have vaccination data in either the CIMS registry or self-reported and were considered unvaccinated. Of almost half of the participants (43.7%) all self-reported vaccination dates fully matched CIMS report. In case dates were the same but vaccine products differed between the two sources, the CIMS registry was leading. Additionally, for 17.8% of the participants vaccination data was almost the same in both sources, with one or more dates less than 2 months apart. In this case, again the CIMS registry was leading for both vaccination date and vaccine product. Thus, in total for 61.5% of the participants only CIMS registry data was used and 2.4% was not vaccinated. Handling of data of the remaining 36.1% of participants is described in the next paragraphs.

Among all participants, 9.2% did not have any vaccination data in the CIMS registry, but did have self-reported vaccination dates. Almost 94% of those did provide informed consent for linking study data and CIMS data. Possible explanations for not having data in CIMS are not being able to link study data and CIMS data, no consent given for vaccination registration in CIMS, or data was not properly registered in CIMS since vaccination was administered by a different organization than the public health service or in a foreign country. Thus, for 9.2% of the participants only self-reported vaccination data was available and used.

In 19.4% of the participants, CIMS registry data appeared incomplete. Dates that were available in both sources corresponded with one another. For example, when comparing the dates, the first dose date in CIMS was similar to the third dose date in the self-reported data suggesting that this participant did receive three doses but the first two were not registered in CIMS. CIMS registry data was obtained in March 2022 while self-reported data was from 4 July 2022, therefore many reported second booster doses were only self-reported and not available in the CIMS registry. In most of the cases where CIMS was incomplete, only booster (15.5%) or second booster dose (78.8%) was missing from the registry data. For this group of participants, CIMS data was used and complemented with self-reported vaccinations. In a much smaller group of participants (3.0%), one or multiple doses were again missing from CIMS, while other dates were not exactly the same between CIMS and self-report but differed a few days. Also in these persons, CIMS dates were used when similar to self-reported data and complemented with self-reported dates of doses missing in CIMS. For a small group of participants (3.1%), doses were reported that were not in CIMS and doses that were in CIMS were not self-reported. Sometimes, some of the vaccination dates did correspond with one another. For example, dose 1 and 2 were the same in CIMS and self-reported data. Yet, dose 3 was self-reported to be in December 2021 and dose 3 in CIMS was in March 2022. In these cases, all data was combined: vaccination 1 and 2 from CIMS, 3 from self-reported data, and 4 from CIMS. In total, for 25.5% of the participants, self-reported vaccination data and registry data was combined, mostly because CIMS registry data did not yet include part of the first and most of the second booster vaccinations.

Lastly, there was a group of participants (1.3%) for whom we were not able to combine dates as they were so different from one another and did not fit into a “normal” vaccination scheme. For these participants, self-reported vaccination data was used.

**Table S1**. Flowchart combining CIMS (until 10 March 2022) and self-reported (until 4 July 2022) vaccination data

| **CIMS registry data?** | | **Self-reported data?** | **CIMS & self-reported data the same?** | **Percent of study population** | **Data used** | **Additional explanation** |
| --- | --- | --- | --- | --- | --- | --- |
| NO | | NO | - | 2.4% | - | Classified as unvaccinated |
| YES | | YES | YES | 43.7% | CIMS |  |
| YES | | YES | <2 months difference | 17.8% | CIMS |  |
| NO | | YES | - | 9.2% | Self-reported data | - No consent linkage CIMS and study data (6.2%) - No consent registration in CIMS - No proper registration in CIMS by organization administering the vaccine |
| INCOMPLETE | | YES | YES | 19.4% | CIMS data complemented with self-reported data | - Most often first or second booster vaccinations missing from CIMS (CIMS was requested in March 2022) |
| INCOMPLETE | | YES | <1 month difference | 3.0% | CIMS data complemented with self-reported data |  |
| INCOMPLETE | | INCOMPLETE | YES | 3.1% | CIMS data complemented with self-reported data |  |
|  | YES | YES | NO | 1.3% | Self-reported data |  |

**Figure S1.** Number of active participants contributing person-time to the analysis* per vaccination status from 12 July 2021 to 6 June 2022

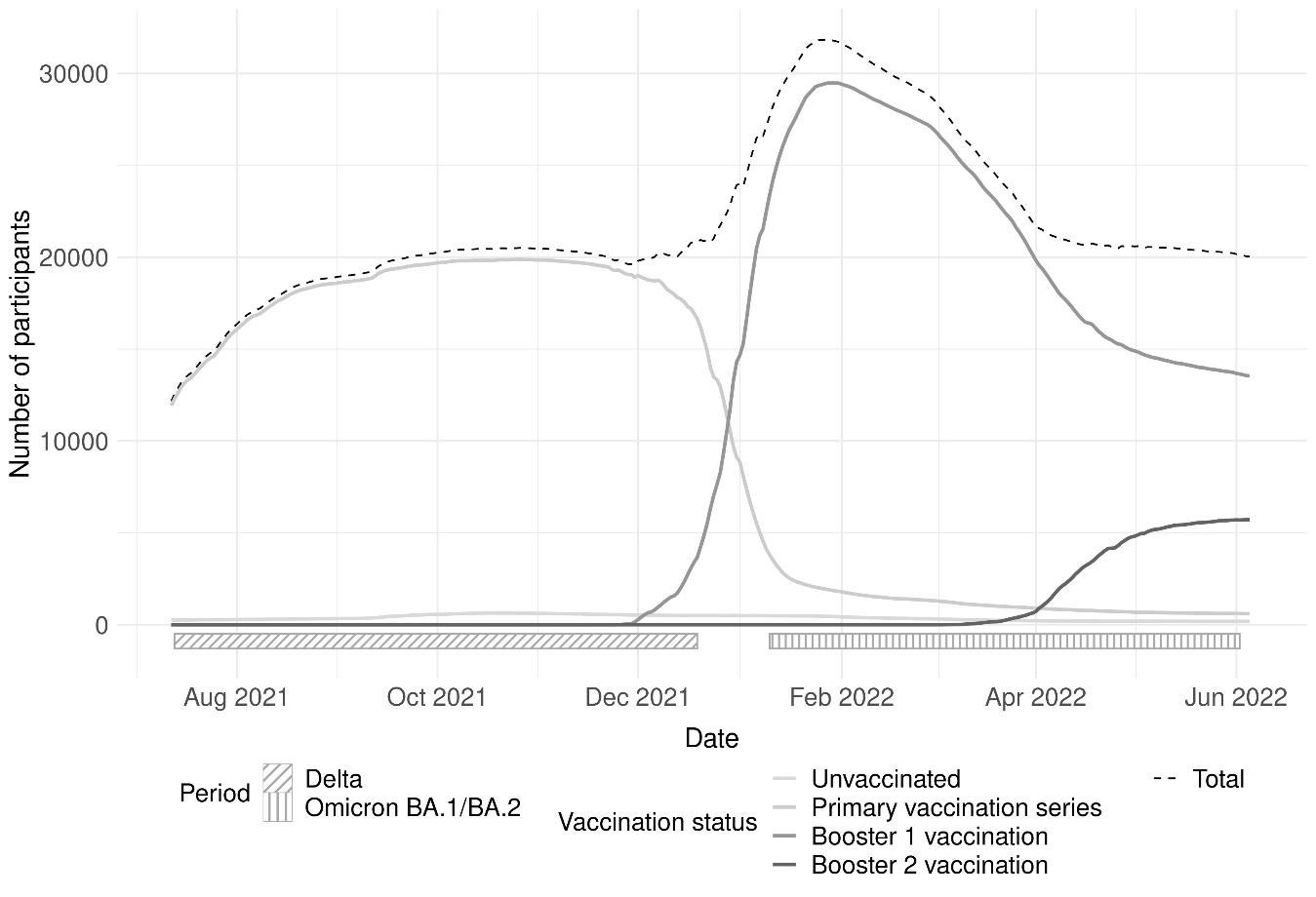

^a^ Participants could have been inactive because of not yet having completed the baseline questionnaire, having an inactive vaccination status (days between vaccine administration and obtained vaccination status), or already being censored.

**Figure S2.** Number of infections reported in VASCO study population by type of test from 12 July 2021 to 6 June 2022

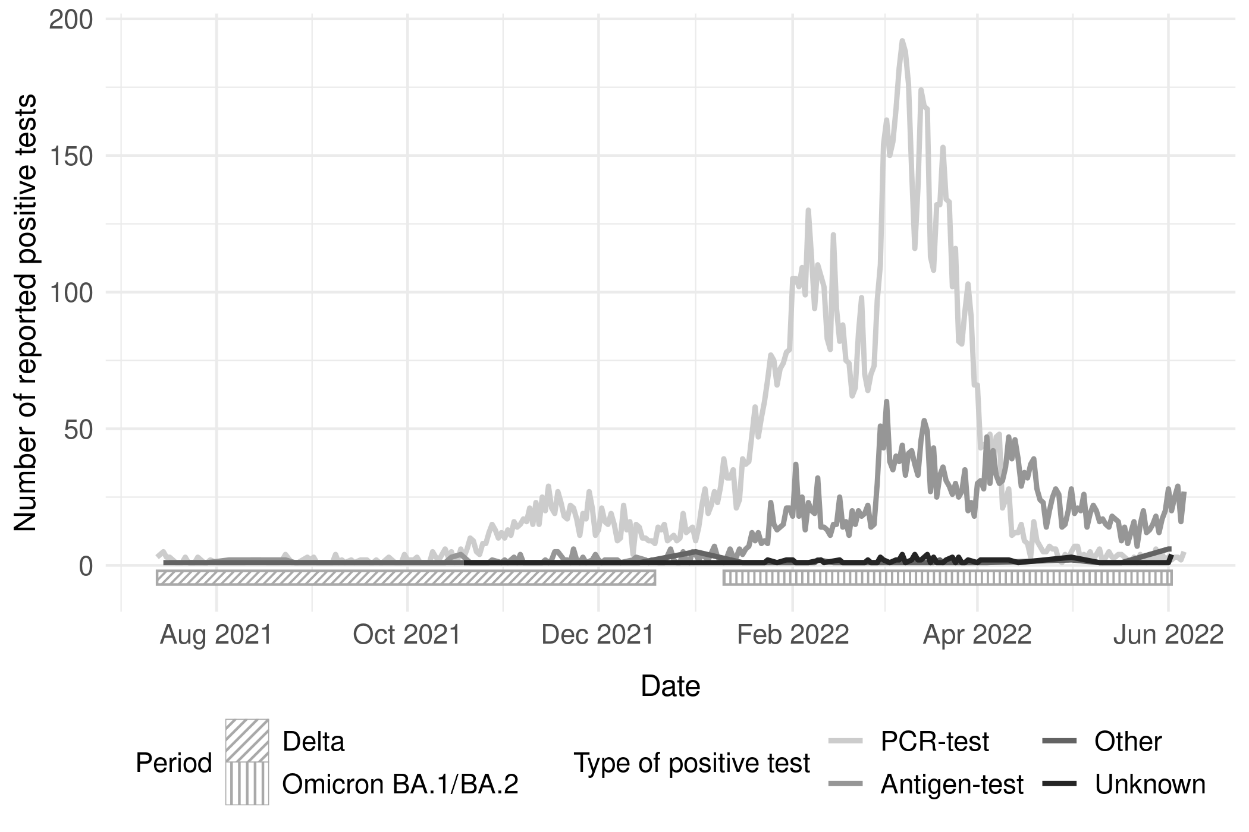

**Figure S3**. Vaccine effectiveness* for primary vaccination series and first booster vaccination in Delta and Omicron BA.1/BA.2 period in participants with high intention to test if experiencing symptoms from 12 July 2021 to 6 June 2022

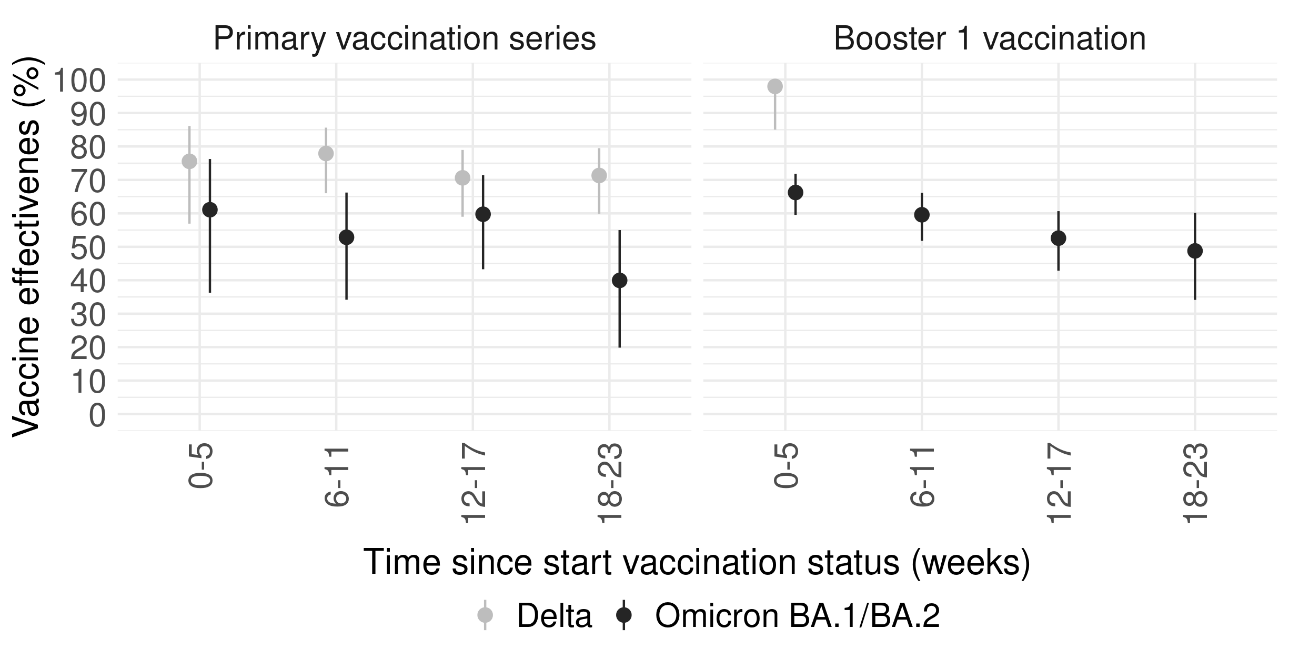

^a^ Adjusted for age group, sex, educational level, medical condition.

**Figure S4.** Intention to test in case of COVID-19-like symptoms from 12 July 2021 to 6 June 2022

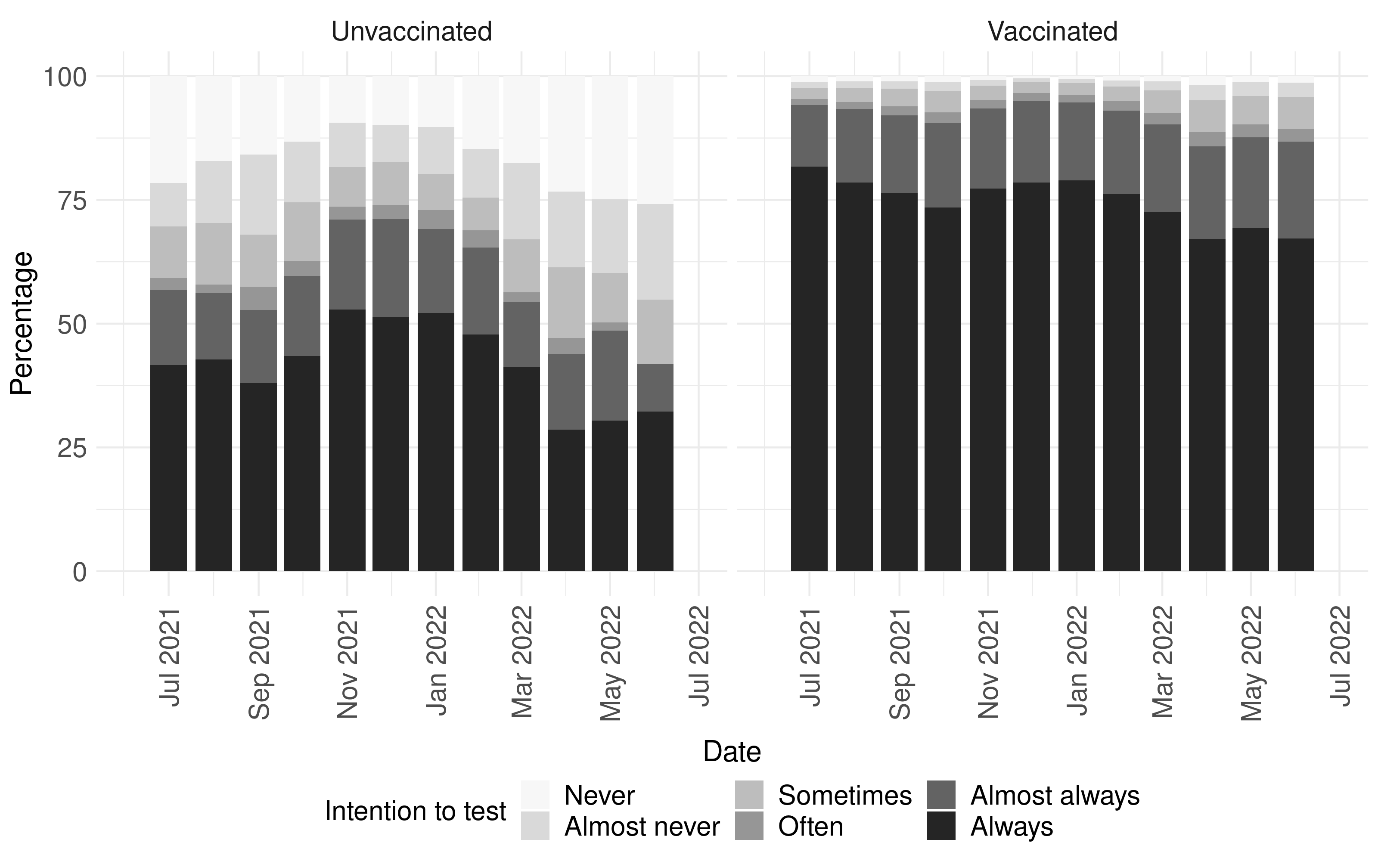

**Table S2.** Vaccine effectiveness^a^ per vaccination status stratified by Delta and Omicron dominant period in participants aged 60 years and older from 12 July 2021 to 6 June 2022

|  | **Number of infections** | **Person-weeks** | **Rate (per 1,000 weeks)** | **Adjusted^1^ VE (95% CI)** |
| --- | --- | --- | --- | --- |
| Delta period |  |  |  |  |
| Unvaccinated | 21 | 3,582 | 5.9 | Ref. |
| Primary series (0-5 weeks) | 19 | 37,252 | 0.5 | 67.3 (19.7-86.7) |
| Primary series (6-11 weeks) | 19 | 64,003 | 0.3 | 79.7 (58.3-90.1) |
| Primary series (12-17 weeks) | 124 | 68,342 | 1.8 | 61.3 (38.0-75.9) |
| Primary series (18-23 weeks) | 263 | 65,101 | 4.0 | 62.1 (40.6-75.8) |
| Booster 1 vaccination (0-5 weeks) | 0 | 2,042 | 0.0 | -- |
| Omicron BA.1-2 period |  |  |  |  |
| Unvaccinated | 78 | 2,509 | 31.1 | Ref. |
| Primary series (0-5 weeks) | 8 | 640 | 12.5 | 45.9 (-12.2-74.0) |
| Primary series (6-11 weeks) | 24 | 902 | 26.6 | 37.5 (1.1-60.4) |
| Primary series (12-17 weeks) | 17 | 1,080 | 15.7 | 63.8 (38.8-78.6) |
| Primary series (18-23 weeks) | 20 | 1,281 | 15.6 | 47.7 (14.4-68.0) |
| Booster 1 vaccination (0-5 weeks) | 148 | 90,840 | 19.9 | 53.7 (41.0-63.8) |
| Booster 1 vaccination (6-11 weeks) | 954 | 94,656 | 10.5 | 48.9 (35.8-59.3) |
| Booster 1 vaccination (12-17 weeks) | 2,757 | 55,787 | 29.1 | 39.3 (22.8-52.3) |
| Booster 1 vaccination (18-23 weeks) | 878 | 26,774 | 15.7 | 40.3 (16.2-57.4) |
| Booster 2 vaccination (0-5 weeks) | 230 | 35,893 | 6.4 | 50.0 (34.0-62.1) |
| Booster 2 vaccination (6-11 weeks) | 40 | 5,495 | 7.3 | 15.6 (-32.5-46.3) |

^a^ Adjusted for age group, sex, educational level, medical condition.

**Table S3.** Vaccine effectiveness^a^ per vaccination status stratified by Delta and Omicron dominant period in participants vaccinated with Comirnaty from 12 July 2021 to 6 June 2022

|  | **Number of infections** | **Person-weeks** | **Rate (per 1,000 weeks)** | **Adjusted^b^ VE (95% CI)** |
| --- | --- | --- | --- | --- |
| Delta period |  |  |  |  |
| Unvaccinated | 126 | 10,500 | 12.0 | Ref. |
| Primary series (0-5 weeks) | 28 | 41,890 | 0.7 | 81.3 (69.3-88.6) |
| Primary series (6-11 weeks) | 57 | 62,500 | 0.9 | 79.3 (71.1-85.2) |
| Primary series (12-17 weeks) | 187 | 65,441 | 2.9 | 73.6 (66.9-79.0) |
| Primary series (18-23 weeks) | 210 | 54,309 | 3.9 | 72.4 (64.9-78.3) |
| Booster 1 vaccination (0-5 weeks) | 1 | 1,365 | 0.7 | 95.5 (67.6-99.4) |
| Omicron BA.1-2 period |  |  |  |  |
| Unvaccinated | 301 | 5,876 | 51.2 | Ref. |
| Primary series (0-5 weeks) | 18 | 450 | 40.0 | - |
| Primary series (6-11 weeks) | 31 | 710 | 43.7 | 45.0 (20.3-62.1) |
| Primary series (12-17 weeks) | 30 | 845 | 35.5 | 43.2 (17.3-61.0) |
| Primary series (18-23 weeks) | 69 | 1911 | 36.1 | 18.0 (-7.3-37.3) |
| Booster 1 vaccination (0-5 weeks) | 687 | 21,469 | 32.0 | 51.2 (43.6-57.8) |
| Booster 1 vaccination (6-11 weeks) | 857 | 19,522 | 43.9 | 45.8 (37.8-52.7) |
| Booster 1 vaccination (12-17 weeks) | 287 | 13,379 | 21.4 | 38.0 (25.7-48.3) |
| Booster 1 vaccination (18-23 weeks) | 70 | 6,050 | 11.6 | 10.5 (-27-36.9) |

^a^ Adjusted for age group, sex, educational level, medical condition.

^b^ VE was not reported when number of person-weeks <500

**Table S4.** Vaccine effectiveness per vaccination status stratified by risk group in the Omicron dominant period from 10 January 2022 to 6 June 2022

|  | **18-59 years with medical risk condition** | **18-59 years without medical risk condition^a^** | **60-85 years with medical risk condition** | **60-85 years without medical risk condition** |
| --- | --- | --- | --- | --- |
| Number of infections | 1,197 | 5,211 | 1,851 | 3,292 |
| Person-weeks | 41,987 | 148,293 | 127,530 | 188,327 |
| Rate (per 1,000 weeks) | 28.5 | 35.1 | 14.5 | 17.5 |
| VE^b^ (95%CI) |  |  |  |  |
| Unvaccinated | Ref. | Ref. | Ref. | Ref. |
| Primary series (0-5 weeks) | - | - | - | - |
| Primary series (6-11 weeks) | - | 53.7 (32.4-68.3) | - | - |
| Primary series (12-17 weeks) | - | 43.7 (18.6-61.1) | 44.9 (-17.9-74.2) | - |
| Primary series (18-23 weeks) | - | 23.1 (1.7-39.8) | 30.2 (-49.4-67.4) | 50.6 (0.5-75.5) |
| Booster 1 vaccination (0-5 weeks) | 40.4 (9.3-60.9) | 58.9 (52.3-64.6) | 43.5 (6.3-65.9) | 56.0 (41.7-66.8)* |
| Booster 1 vaccination (6-11 weeks) | 27.5 (-9.6-52.1) | 51.3 (43.5-58.0) | 30.3 (-13.0-56.9)* | 54.2 (40.7-64.7) |
| Booster 1 vaccination (12-17 weeks) | 19.7 (-25.1-48.5) | 44.0 (33.7-52.7) | 16.2 (-38.2-49.2)* | 46.3 (29.2-59.3)* |
| Booster 1 vaccination (18-23 weeks) | 3.7 (-74.8-47.0)* | 34.3 (14-49.8) | 26.4 (-41.2-61.6) | 44.1 (16.6-62.6) |
| Booster 2 vaccination (0-5 weeks) | - | - | 20.0 (-37.9-53.6) | 59.4 (43.5-70.9) |
| Booster 2 vaccination (6-11 weeks) | - | - | 4.8 (-114.2-57.7) | 14.9 (-47.6-51.0) |

^a^ Reference group for interaction

^b^ VE was not reported when number of person-weeks <500

* p-value interaction vaccination status and medical risk group < 0.05

**Table S5.** Vaccine effectiveness^a^ of primary vaccination series stratified by vaccine product in the Delta and Omicron dominant period from 12 July 2021 to 6 June 2022

|  | **Adjusted^b^ VE (95% CI) Comirnaty** | **Adjusted^b^ VE (95% CI) Spikevax** | **Adjusted^b^ VE (95% CI) Vaxzevria** | **Adjusted^b^ VE (95% CI) Jcovden** |
| --- | --- | --- | --- | --- |
| Delta period |  |  |  |  |
| Unvaccinated | Ref. | Ref. | Ref. | Ref. |
| Primary series (0-5 weeks) | 81.3 (69.2-88.6) | 100 (-Inf-100) | 68.0 (12.3-88.3) | 58.5 (-14.9-85) |
| Primary series (6-11 weeks) | 79.3 (71.1-85.2) | 83.7 (61.4-93.1) | 77.2 (47.6-90.1) | 56.6 (-7.4-82.5) |
| Primary series (12-17 weeks) | 73.6 (66.9-79.0) | 77.2 (65.7-84.8) | 58.0 (40.2-70.4) | 69.6 (48.5-82.1) |
| Primary series (18-23 weeks) | 72.4 (64.9-78.3) | 87.6 (78.1-93.0) | 65.3 (52.9-74.5) | 75.0 (62.3-83.5) |
| Omicron BA.1-2 period |  |  |  |  |
| Unvaccinated | Ref. | Ref. | Ref. | Ref. |
| Primary series (0-5 weeks) | - | - | - | - |
| Primary series (6-11 weeks) | 45.0 (20.3-62.1) | - | - | - |
| Primary series (12-17 weeks) | 43.2 (17.3-61) | - | - | - |
| Primary series (18-23 weeks) | 18.0 (-7.3-37.3) | 15.8 (-26.3-43.9) | - | - |

^a^ VE was not reported when number of person-weeks <500

^b^ Adjusted for age group, sex, educational level, medical condition.

**Table S6.** Vaccine effectiveness^a^ of first booster vaccination stratified by vaccine product in the Delta and Omicron dominant period from 12 July 2021 to 6 June 2022

|  | **Adjusted^b^ VE (95% CI) Comirnaty** | | | **Adjusted^b^ VE (95% CI) Spikevax** | | |
| --- | --- | --- | --- | --- | --- | --- |
|  | **Any primary vaccination series** | **mRNA primary vaccination series** | **Vaxzevria primary vaccination series** | **Any primary vaccination series** | **mRNA primary vaccination series** | **Vaxzevria primary vaccination series** |
| Delta period |  |  |  |  |  |  |
| Unvaccinated | Ref. | Ref. | Ref. | Ref. |  | Ref. |
| Booster 1 vaccination (0-5 weeks) | 94.3 (81.9-98.2) | 97.4 (81.2-99.6) | 90.9 (34-98.8) | 100 (-Inf-100) | 100 (-Inf-100) | - |
| Booster 1 vaccination (6-11 weeks) | - | - | - | - | - | - |
| Booster 1 vaccination (12-17 weeks) | - | - | - | - | - | - |
| Booster 1 vaccination (18-23 weeks) | - | - | - | - | - | - |
| Omicron BA.1-2 period |  |  |  |  |  |  |
| Unvaccinated | Ref. | Ref. | Ref. | Ref. |  | Ref. |
| Booster 1 vaccination (0-5 weeks) | 51.8 (45.2-57.7) | 51.1 (44.1-57.3) | 58.4 (48.8-66.1) | 66.1 (61-70.5) | 68.3 (63.1-72.8) | 67.6 (60.1-73.7) |
| Booster 1 vaccination (6-11 weeks) | 45.5 (38.3-51.8) | 45.2 (37.7-51.8) | 44.5 (34.5-53) | 53.0 (46.5-58.7) | 56.9 (50.5-62.5) | 45.3 (34.0-54.8) |
| Booster 1 vaccination (12-17 weeks) | 35.8 (26.1-44.2) | 40.3 (30.4-48.8) | 21.6 (3.4-36.3) | 38.9 (28.3-47.9) | 34.8 (21.3-46) | 37.1 (19.6-50.9) |
| Booster 1 vaccination (18-23 weeks) | 19.1 (-3.1-36.5) | 25.4 (2.1-43.2) | -2.6 (-68-37.3) | 32.6 (9.8-49.7) | 23.4 (-10.1-46.8) | 47.1 (12.4-68.1) |

^a^ VE was not reported when number of person-weeks <500

^b^ Adjusted for age group, sex, educational level, medical condition.
